# Scientists’ opinion, attitudes, and consensus towards immunity passports

**DOI:** 10.1101/2021.02.02.21250796

**Authors:** Iván Aranzales, Ho Fai Chan, Reiner Eichenberger, Rainer Hegselmann, David Stadelmann, Benno Torgler

**Affiliations:** Centre for Behavioural Economics, Society and Technology (BEST), Brisbane, Queensland, Australia; CREMA - Center for Research in Economics, Management and the Arts; Frankfurt School of Finance & Management, Germany; IREF - Institute for Research in Economic and Fiscal Issues; School of Economics and Finance, Queensland University of Technology, Brisbane, Queensland, Australia; University of Bayreuth, Germany; University of Fribourg, Switzerland

## Abstract

**Objectives:** We measured attitudes towards “immunity passports” in the context of COVID-19 of a large sample of scientists. Consensus of scientists’ opinions on a different aspect of immunity passports was assessed.

**Methods:** We designed and implemented a survey to capture what scientists from around the world and different scientific background think about immunity certification. The survey was sent to the corresponding authors of scholarly articles published in the last five years in the top 20-ranked journals in each of the 27 subject areas between May and June 2020. Responses from 12,738 scientists were captured, and their distribution was tabulated by participants in health science and other fields. Consensus of responses was calculated using a variant of Shannon Entropy, made suitable for the ordinal response variables.

**Results:** Half of the scientists surveyed, regardless of academic background agree that a potential immunity passport program will be good for public health (50.2%) and the economy (54.4%), with 19.1% and 15.4% of participants disagree, respectively. A significant proportion of scientists raised concerns about immunity certification over fairness to others (36.5%) and social inequality (45.5%). There is little consensus in the different aspects of immunity passport among scientists. Overall, scientists with health background hold a more conservative view towards immunity certification.

**Conclusions:** Our findings suggest a lack of general agreement regarding the potential health and economic benefits, societal costs, and ethical issues of an immunity certification program within the scientific community. Given the relevant and important implications of immunity passport due to the increasing vaccine availability and efficacy, more attention should be given to the discussion of the design and implementation of immunity certification program.

**Strengths and limitations of this study:**

- First cross-disciplinary survey with a large and international sample size that enables mapping of scientists’ opinions and attitudes towards COVID-19 immunity certificates.
- From the survey responses, we measured, reported, and compared the levels of consensus of scientists between health-related and non-health-related discipline.
- Response rate and sample representativeness are moderate.

## INTRODUCTION

We have reached the roll-out stage of COVID-19 vaccines. The United Kingdom was the first country to approve a vaccine tested through large clinical trials; on December 8, 2020, the government started administering the Pfizer and BioNTech vaccine. This makes the potential introduction of immunity based-licenses (“immunity passports” or “immunity certificates”) an important policy topic requiring further analysis, particularly with respect to its acceptance, given increasing evidence^1^ showing the effectiveness of antibodies from natural-^2^ and vaccine-induced^3–4^ can help protect against SARS-CoV-2 infection. Scholarly debates on the benefits of and objections to immunity certificates are ongoing and without clear consensus^5–9^. However, due to the uniform and controlled nature of the treatment, the advantage of vaccine-induced immunity is that patterns and duration of immunity are more predictable than infection^5^. As scientists play an vital role in the immunity certification debate, we present evidence from a large-scale survey of 12,738 scientists (of which 4,852 are health scientists).

## METHODS

### Survey data collection

Responses were gathered via the SurveyMonkey platform from scholars appearing in Scopus who have published in the top 20 journals of each subject areas listed in SCImago Journal Rank in the last five years (including also scholars from the bibliographic database RePEc to increase the representation of social scientists). Data were collected between May 4 and June 3, 2020 from a sample of 213,648 emails. The response rate was 14% based on emails opened. Ethical approval for the survey and the data collection was given on April 23, 2020 by the Ethics Commission of the Frankfurt School of Finance & Management (Frankfurt, Germany).

### Statistical analyses

Statistical analyses were conducted using STATA (MP 16.1) statistical software. The two-sample Wilcoxon rank-sum test was used to assess the statistically significant differences (two-tailed test) in the responses between participants from health sciences and other disciplines.

Consensus of the ordinal response variable *X* with *i* categories is defined as: 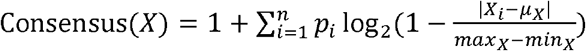, where *p* is the share of responses (excluding non-responses)^10,11^. A value of 0 indicates the participants’ responses are evenly split to the two extremes, while a value of 1 means that all responses are in the same category. The consensus score is around 0.45 (depending on the number of response categories) if responses are evenly split into each category. 95% confidence intervals (error bars) of the consensus measure are constructed by performing bootstrap resampling with 300 replications. The two-sample *t*-test was used to test for statistically significant differences (two-tailed) in the consensus scores between health and non-health related participants. Computing consensus using the Shannon Entropy equation, i.e., 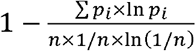, yields identical qualitative conclusions.

### Patient and Public Involvement

No patients were involved in the development of the research question, design and implementation of the study, or interpretation of the results.

## RESULTS

About half of scientists agree (to varying degrees) that the issue of immunity certificates for the duration of immunity is good for public health (Figure 1a) and the economy (Figure 1b), while one-fifth and one-sixth disagree, respectively. Yet, in terms of fairness, about 40% (health sciences) and 35% (other disciplines) of scholars think that issuing immunity certificates will *not* be fair to those who do not have immunity (Figure 1c). Around 45% of the respondents think that immunity certificates will increase inequality in society (Figure 1d). Relative to other scholars, health scientists are less in favour of immunity certificates for the perceived benefit to public health (*P*=.0006) and the economy (*P*=.0091) or fairness (*P*<.0001) and equality concerns (*P*=.057). Overall, 38.55% and 44.4% participants indicate that they pay (a positive amount) for the opportunity to obtain an immunity certificate that will allow the easing of social-isolation and travel restrictions, respectively. A relatively larger proportion of health scientists (compared to all other disciplines) say that they will not pay to have an immunity certificate, and are less likely to pay a larger amount for an immunity passport (Figure 1e: *P*<.0001; Figure 1f: *P*<.0001).

**Figure 1.**
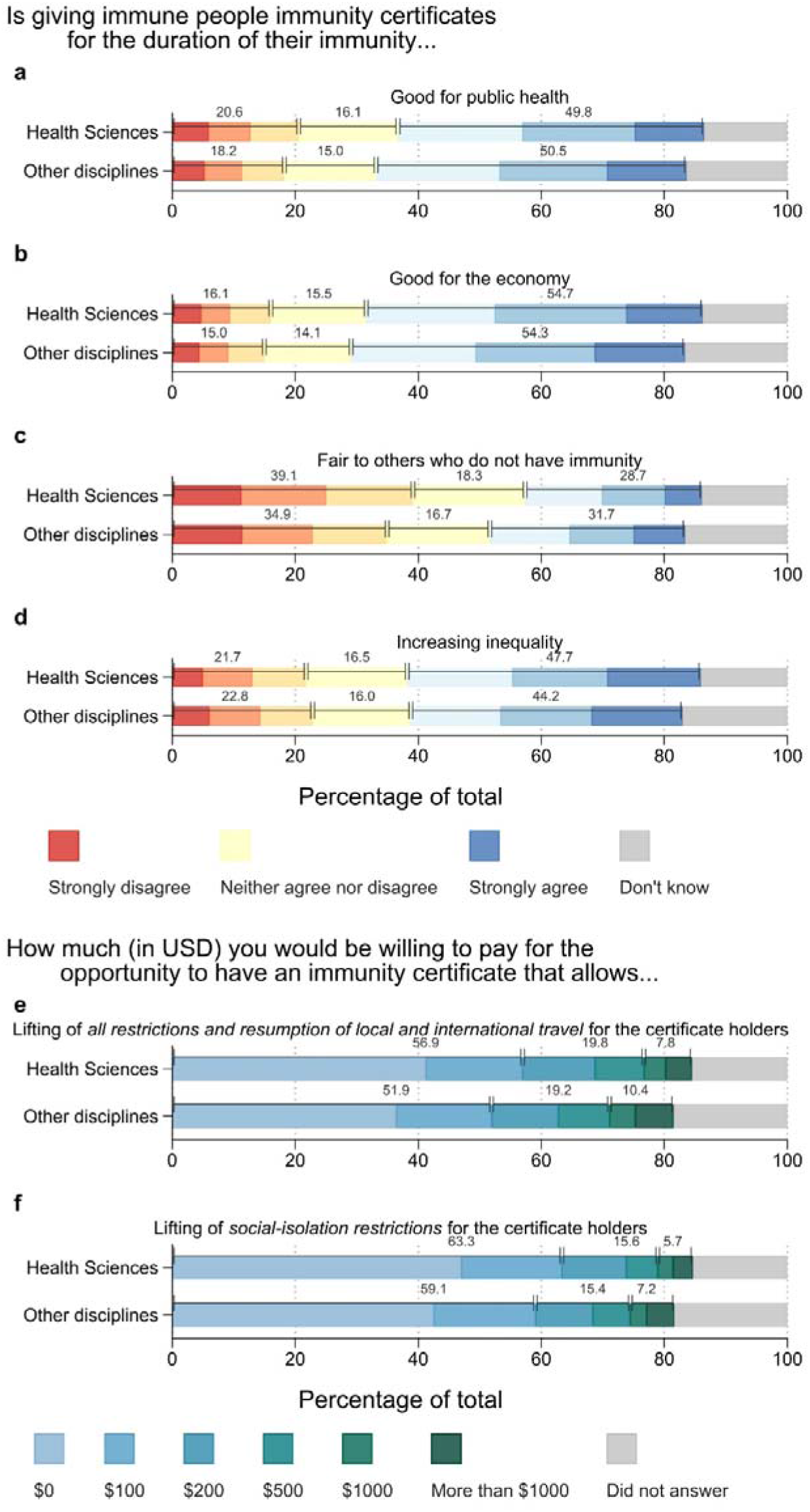
Perceptions of Immunity Passports and Willingness to Pay for Immunity Passports. *Notes*: N = 12,738. Health sciences: *n* = 4,852, median age group 40 to 44; 51.6% female; other disciplines: *n* = 7,886, median age group 40 to 44; 36.2% female). Figure 1 shows the distribution of responses to the question “Is giving immune people immunity certificates for the duration of their immunity…” **a)** *good for public health*; **b)** *good for the economy*; **c)** *fair to others who do not have immunity*; **d)** *Increasing inequality* (left) and participants’ self-report willingness to pay to receive an immunity certificate (**e** and **f**), by field of research. For panel **a** to **d**, numbers above the bars represent the percentage of all participants reporting a value of 1 to 3 (disagree to some extent), 4 (neutral), and 5-7 (agree to some extent).

Overall, there is no general agreement; out of the four opinions on immunity certificates, concerns for fairness and inequality have lower consensus compared with public health and the economy (Figure 2a). While the level of agreement does not statistically differ between health and other scientists with respect to public health (*P*=.920) and the economy (*P*=.283), health scientists have relatively higher consensus on concerns of fairness (*P*=.0001), and equality (*P*=.0007). Additionally, health scientists report a stronger consensus regarding the willingness to pay for an immunity certificate (Figure 2b: easing of social isolation restrictions (*P*=.0001); lifting of all restrictions including travel (*P*<.0001)).

**Figure 2.**
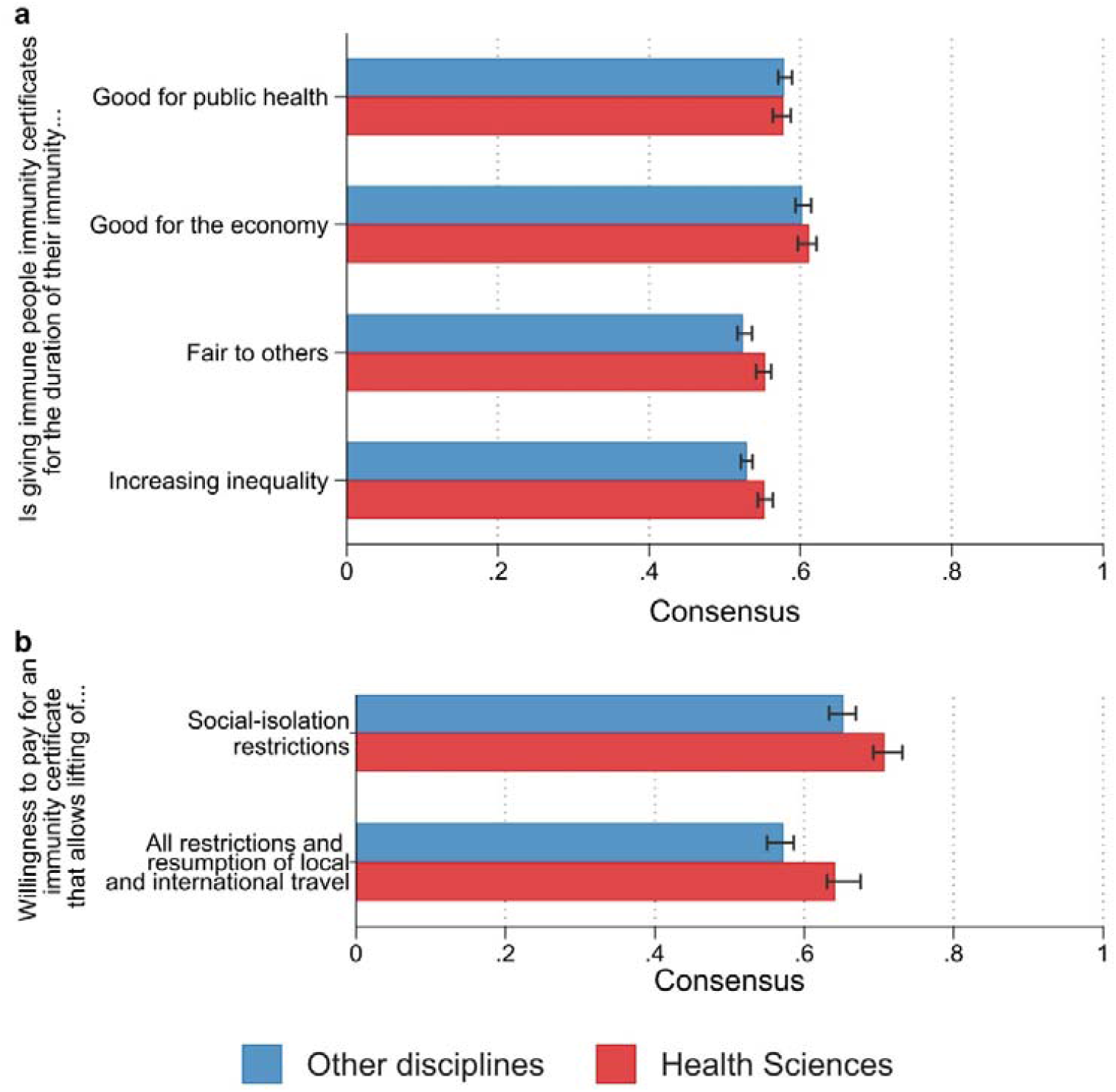
Opinion Consensus on Immunity Passports. *Notes*: Figure 2 shows the levels of agreement among participants’ responses by field of study (health sciences and other disciplines).

## DISCUSSION

As our survey on scientists’ perceptions of immunity certificates was conducted when vaccinations were not yet available, our results can be interpreted as conservative evidence on the acceptability of vaccination certificates. Health scholars are less supportive of immunity certificates; the reasons for this require a more detailed discussion as their policy influence and voice during a pandemic are essential in formulating solutions. Policies can be designed to address issues such as inequality or fairness, and such concerns may decrease once more people are vaccinated^12–14^. Moreover, convalescent immunity certificates should not be ignored. It is expected that one-third of the world’s population will have access to a COVID-19 vaccine by the end of 2021, however many people in lower-income countries might have to wait until 2023 or 2024 for vaccination^15^. Canada, the US, the UK, Australia and the EU have already ordered about half the expected capacity for 2021. Developing countries are particularly vulnerable to the economic effects of lockdown and social distancing measures. In addition, understanding the use of immunity passports is also useful in planning our response to future infectious diseases.

## Data Availability

Data and codes used in this study are accessible via OSF.

https://osf.io/xghq7/

## Author Contributions

Mr Aranzales and Drs Chan, Stadelmann, and Torgler had full access to all of the data in the study and take responsibility for the integrity of the data and the accuracy of the data analysis.

*Concept and design:* Eichenberger, Hegselmann, Stadelmann, Torgler.

*Acquisition, analysis, or interpretation of data*: Aranzales, Chan, Stadelmann, Torgler.

*Drafting of the manuscript:* Aranzales Chan, Stadelmann, Torgler.

*Critical revision of the manuscript for important intellectual content:* All authors.

*Statistical analysis:* Chan.

*Supervision:* Chan, Stadelmann, Torgler.

